# Concordance Between a Temple-Worn Optical Wearable and Transcranial Doppler During Exercise and Postural Transitions in Healthy Adults

**DOI:** 10.64898/2026.09.02.26362022

**Authors:** Anit Kumar, Laura van Rosmalen, Anushka Gupta, Shubham Kumar Sharma, Ramesh Chandra Gupta, Satchidananda Panda, Neelu Jain Gupta

**Author notes:** These authors contributed equally to this work (Anit Kumar, Laura van Rosmalen).

## Abstract

Cerebral hemodynamics are difficult to monitor continuously outside the laboratory. Optical head-worn wearables have been proposed for tracking cerebral blood-flow signals, but they require comparison with an established cerebrovascular reference before they can be interpreted. We evaluated a temple-worn optical wearable, Temple, that outputs a proprietary, dimensionless Brain Flow index, intended as a proxy for relative changes in cerebral hemodynamics, against transcranial Doppler (TCD) ultrasound, which measures blood-flow velocity in the middle cerebral artery (MCAv). Twenty-three healthy adults completed two physiological challenges that elicit distinct and acute cerebral hemodynamic responses: a cycle-ergometer exercise protocol and a stand-to-supine postural transition protocol. Twenty participants were analyzed per protocol. The Brain Flow index tracked MCAv in both protocols, with significant within-subject temporal correlations (median Pearson r = 0.795 and 0.799 for exercise and postural transition; p < 0.001) and directionally concordant, statistically significant transition responses for both increases and decreases in flow. Bland-Altman analysis of the normalized transition responses showed small mean biases between the two devices, consistent with similar relative response shapes. Because both signals were standardized within session before this comparison, it addresses the shape of the relative change rather than agreement in absolute units. The Brain Flow index reproduced the direction and time course of MCAv under both perturbations, including the postural transition, where heart rate moved in the opposite direction. Further studies using complementary modalities and additional cerebrovascular reactivity challenges are required to establish clinical use cases and cerebral specificity of the Brain Flow index.

## 1. Introduction

The brain has minimal intrinsic energy reserves and therefore depends on a continuous, tightly regulated blood supply to meet its metabolic demands [1-3]. Cerebral blood flow is dynamically adjusted to maintain adequate perfusion during different behavioral, cardiovascular, and environmental conditions. Some adjustments occur acutely, such as during physical activity [4,5] or changes in posture [6,7], whereas longer-term influences, including sleep [8-10], diet [11], stress [12], caffeine [13], smoking [14], and alcohol use [15-17], may shape cerebrovascular health over extended periods. This sensitivity has made cerebral hemodynamics an informative marker of vascular brain health, with relevance to aging, cognition, and cerebrovascular risks [18].

Despite this importance, cerebral perfusion remains difficult to measure outside clinical or research environments. Perfusion imaging methods such as Positron Emission Tomography (PET), Single-Photon Emission Computed Tomography (SPECT), xenon-enhanced Computed Tomography (CT), and MRI-based approaches, including arterial spin labeling, provide detailed spatial maps of cerebral perfusion but require a fixed scanner and a stationary participant and are poorly suited to repeated monitoring during everyday physiological challenges [19]. Wearable optical sensors could extend monitoring into these settings, but because they measure light at the surface of the head, their signals also reflect scalp and systemic circulation [20]. As a result, they cannot be assumed to index brain hemodynamics until they are validated against an established cerebrovascular reference.

Transcranial Doppler (TCD) ultrasound provides an appropriate reference for this comparison. It enables continuous, high-temporal-resolution assessment of blood-flow velocity in the large basal cerebral arteries, most commonly the middle cerebral artery (MCA), and is well suited to characterize rapid cerebrovascular responses during perturbations such as exercise, orthostatic stress, and postural change [21,22]. MCA velocity (MCAv) responses to these perturbations have been extensively characterized and MCAv provides a reference signal for evaluating whether a new measurement approach can detect similar changes. TCD has also previously been used to evaluate novel devices designed to measure comparable physiological responses [23].

Arterial pressure and other systemic variables can be treated as inputs that the cerebral circulation transforms into a flow response. Dynamic cerebral autoregulation is defined as the frequency-dependent relationship between mean arterial pressure (MAP) and CBF or CBF velocity, commonly characterized by transfer-function gain, phase, and coherence [24,25]. Posture and exercise perturb this system through different combinations of MAP, venous return, cardiac output, arterial CO2, autonomic activity, and metabolism [4-7]. Because posture and exercise engage these inputs in different combinations, testing a wearable signal against MCAv across both perturbations is a stronger design than testing it during a single manoeuvre.

The device evaluated here, referred to as Temple, is an optical wearable worn on the anterior temporal region, superior to the zygomatic arch and lateral to the lateral orbital rim. It outputs the Brain Flow index, a proprietary signal intended to reflect blood flow to the brain during normal daily activities. The anterior temple is an accessible site for such a wearable: it carries superficial cranial vasculature (including branches of the superficial temporal artery) and lies near peri-orbital networks with anastomotic connections to the internal carotid circulation [26,27]. Because the sensor sits at the surface of the head and reports a composite index rather than a direct measure of cerebral blood flow, its physiological relevance must be carefully validated. We evaluated the final output of the Temple device against simultaneously recorded TCD-derived measurements of cerebral blood flow velocity to assess whether the Brain Flow index responds to physiological protocols in a manner consistent with expected cerebrovascular behavior.

To evaluate whether the Brain Flow index captures physiologically meaningful changes, we used two protocols designed to produce distinct cerebrovascular responses. The exercise protocol was selected to elicit an exercise-related increase in cardiovascular demand, as cycling increases heart rate and, in prior TCD studies, increases MCAv [28-30]. The postural transition protocol was selected to probe responses to changes in body position: although heart rate generally decreases when moving from standing to supine, MCAv is lower during standing and increases during supine rest, consistent with improved cerebral perfusion when supine [6,7,31,32]. These posture-related differences also align with a more recent view of cerebral autoregulation, in which cerebral blood flow is not held constant across a wide range of arterial pressure, as classically proposed, but is instead modulated by the relatively small and often rapid changes in perfusion pressure encountered in everyday life [33,34]. These two protocols have distinctive and well-characterized hemodynamic signatures [6,7,28-31]; they place heart rate and cerebral blood flow velocity in different relationships, making them informative tests of a cerebrovascular claim. Together, they provided complementary models for assessing whether the Brain Flow index tracks directionally consistent changes in cerebral hemodynamic activity relative to TCD-derived MCAv.

## 2. Materials and Methods

### 2.1. Participants

All protocols were conducted in accordance with the Declaration of Helsinki, as revised in 2024 [35], and with the recommendations and standard operating procedures of the Department of Health Research (DHR) and Indian Council of Medical Research (ICMR), India, including voluntary participation and financial compensation as per the guidelines. Protocols were approved by the Institutional Human Ethics Committee of Lala Lajpat Rai Memorial Medical College, Chaudhary Charan Singh University, Meerut, India. This was a non-invasive observational method-comparison study in healthy volunteers rather than a clinical trial of an intervention, and ethics approval was obtained before recruitment began.

Volunteers were eligible if they were aged 18-30 years, had no cardiovascular, pulmonary, metabolic, or neurological disease, had no recent hospitalization, and were taking no medications. Female volunteers were additionally required to be non-pregnant, confirmed by self-report at screening. Participants were instructed to abstain from alcohol and caffeine for at least 12 h before testing, as both acutely alter cerebral perfusion and MCAv [13,16], and to consume their last meal at least 2 h before the session. Participants reporting dizziness or discomfort during a protocol or during data breaks were withdrawn.

Recruitment and data collection took place in February 2026. Of 48 volunteers who registered, 32 met the eligibility criteria and were enrolled, and 23 completed both protocols. After exclusions for recording quality, applied independently within each protocol (Section 2.4), 20 participants were analyzed in the exercise protocol and 20 in the postural transition protocol; because exclusions were protocol-specific, these two samples overlap but are not identical. 17 participants contributed to both analyses, 3 to the exercise analysis only, and 3 to the postural transition analysis only, so 23 individuals contributed to at least one protocol.

The 23 participants who completed both protocols (15 male, 8 female) had a mean age of (23.13 ± 0.5) years and a body mass index of (25.78 ± 1.06) kg/m^2^. They spanned a range of skin tones (Monk Skin Tone MST 2 to MST 8 [36]); skin tone was recorded as a potential moderator of optical sensor performance [37].

Recording sessions were scheduled between 08:00 and 20:00 and conducted in a temperature-controlled environment maintained at approximately 25-30 °C. Neither time of day nor room temperature was standardized further; because skin blood flow contributes to a surface optical signal and is temperature-sensitive, and because cerebral blood flow varies across the day, both remained uncontrolled sources of between-session variability.

### 2.2. Data Acquisition and Devices

During all protocols, participants were monitored simultaneously with a single TCD ultrasound probe placed using a fitted headframe (Figure 1a) over the transtemporal window, and the Temple device was placed on the contralateral temple area (Figure 1b). Unilateral MCAv (cm/s) was measured using non-imaging (blind) pulsed-wave TCD with a 2 MHz monitoring probe (Dolphin IQ, Viasonix, Ra’anana, Israel), fixed over the temporal window with an adjustable headband and ultrasound gel. Time-averaged mean blood-flow velocity was acquired at 1 Hz, with the raw signal sampled at 125 Hz (Figure 1c). The MCA was identified at the start of each session by insonation depth, flow direction, and waveform characteristics, and the probe was then locked in the headframe so that insonation depth and angle were held constant within a participant. No angle correction was applied, as is standard for MCA monitoring through the transtemporal window, and the probe was not reacquired or re-adjusted per protocol.

**Figure 1.**
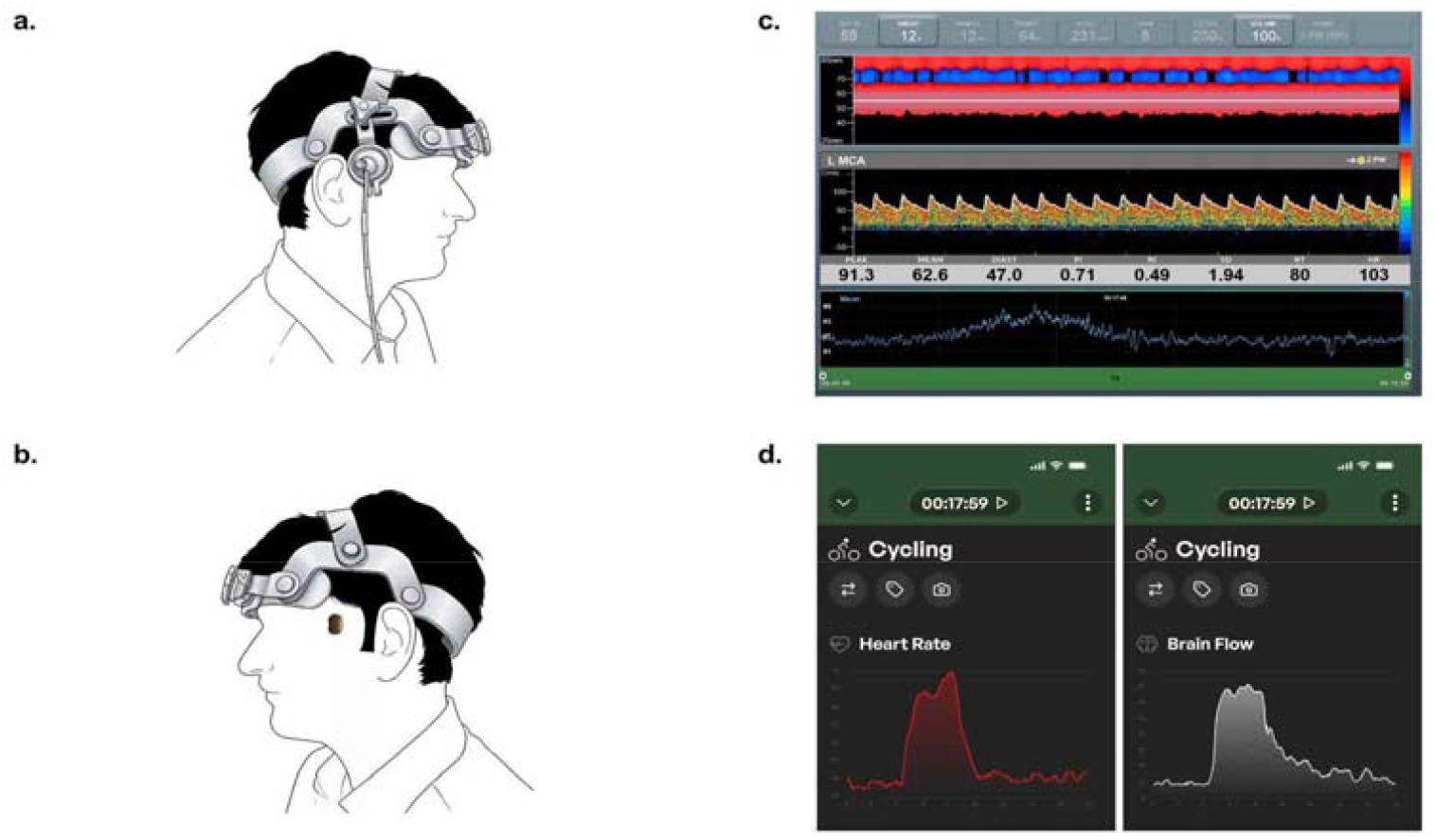
Illustration of the TCD and Temple devices and their respective software interfaces. **(a)** Placement of the transcranial Doppler (TCD) probe over the temporal acoustic window using a headframe. **(b)** Participants simultaneously placed the Temple device on the contralateral temple, without touching the headframe. **(c)** Representative screenshot of the TCD acquisition software displaying middle cerebral artery blood-flow velocity signals and related parameters. **(d)** Representative screenshots of the Temple mobile application displaying heart rate and Brain Flow index measurements.

The Temple device is a continuous-wear, multi-wavelength (green, red, and infrared) reflectance photoplethysmography (PPG) sensor placed over the anterior temple and secured with a medical-grade double-sided adhesive grip. Data were collected in workout mode at 1 Hz through the Temple app, ver. 9.6 of the Brain Flow algorithm (Figure 1d). The device outputs the ‘Brain Flow index’ (Temple-BF), a relative, dimensionless measure of hemodynamic change normalized to each participant’s baseline; it is described as a composite of three families of optical features drawn from the three wavelengths: PPG-based perfusion features, hemoglobin-related optical changes, and cardiac pulse-waveform morphology. Because these wavelengths differ in their tissue penetration, with green light returning predominantly from the most superficial vascular bed and red and infrared light returning from progressively deeper tissue, the composite draws on more than one depth of optical sampling. Because the algorithm is proprietary and closed-source, we did not examine its internal components and evaluated the device solely based on its inputs and outputs.

To preserve independence, the manufacturer was blinded to participant identity, study protocol, and reference measurements. It received only de-identified session timestamps and returned the corresponding Brain Flow index and heart rate values. The pipeline that produced the Brain Flow index had no access to task labels, epoch boundaries, TCD values, or any study outcome. The algorithm version was fixed at 9.6 for the duration of the study.

### 2.3. Protocol Design

Each participant completed two protocols in the same session: an exercise protocol followed by a postural transition protocol (Figure 2). The exercise protocol comprised three consecutive blocks: 240 s of seated baseline rest on a cycle-ergometer, 240 s of fast seated cycling at the participant’s maximum self-selected cadence, and 600 s of seated recovery. After a 30-min break, participants completed the postural transition protocol, four 240s blocks in the order standing, supine, standing, and supine. During the standing blocks, participants stood with their backs against a wall. Standing is a standard orthostatic challenge [38]. Back support was used to standardize the standing blocks and to limit leg-muscle activity, which contributes to venous return during unsupported standing [39]. In all protocols, participants were asked to remain as still as possible during each block and to avoid any unnecessary body movements, including speaking, jaw, and eyebrow movements.

**Figure 2.**
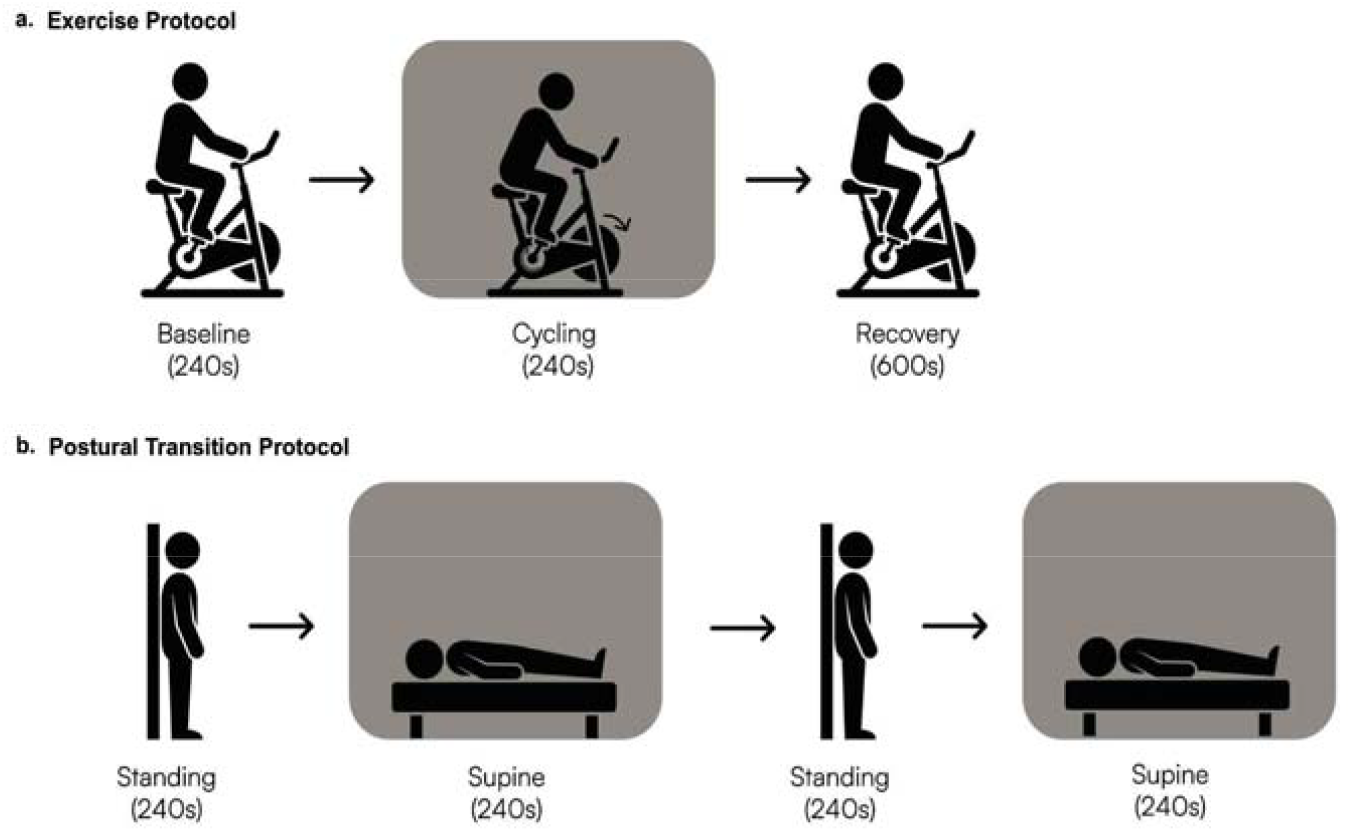
Experimental protocol (schematic). Each participant completed (**a**) an exercise protocol and, after a 30-min break, (**b**) a postural transition protocol. Intervention blocks (cycling; supine) are shaded; block durations are indicated.

### 2.4. Data Analysis

TCD-derived MCAv and heart rate data were exported as CSV files using Viasonix software. Brain Flow index and heart rate data were obtained in CSV format from Temple’s team.

In some instances, poor-quality TCD recording occurred when the headframe used to position the TCD probe moved during the session. The Temple device did not contact the headframe (Figure 1b), but a head or trunk movement large enough to displace the headframe could also disturb the adhesive-mounted optical sensor at the same moment, so a single movement could degrade both recordings. Recordings were screened for signal loss and movement artifacts before any concordance analysis was run, and the device whose trace was degraded determined the reason for exclusion. On these grounds, three participants were excluded from each protocol, in each case two for noisy TCD recordings and one for a noisy Temple recording, leaving 20 analyzable datasets per protocol. Only the affected protocol was excluded for each participant, not the participant’s full dataset. Because no comparisons are made across protocols, the differences in the analyzed samples do not affect the analyses.

All analyses were performed in Python (version 3.13.11) using the SciPy and NumPy libraries [40,41]. MCAv and Brain Flow index time series were analyzed separately for each protocol. TCD reports MCAv in cm/s, whereas the Brain Flow index is a dimensionless proprietary index with no physical unit, so the two cannot be compared on a common measurement scale. Both signals were therefore standardized within the session so that the relative change produced by each protocol could be compared directly. For each participant, device, and protocol, the complete trace was normalized using a z-score transformation based on the mean and standard deviation of the full recording session. The median z-score was then calculated within each task epoch. Transition responses were defined as the change in median z-score between consecutive epochs. For the exercise protocol, these were the baseline-to-cycling and cycling-to-recovery transitions; for the postural transition protocol, the standing-to-supine response was the mean of the two standing-to-supine transitions, and the supine-to-standing response was taken from the single supine-to-standing transition. For each device and transition, subject-level responses were summarized across participants as the median, with 95% confidence intervals estimated by percentile bootstrapping.

### 2.5. Statistical Analysis

An a priori sample size was computed in G*Power 3.1 [42] for a bivariate-normal correlation test. With a two-tailed α = 0.05, power (1 - β) = 0.90, and an expected large effect (coefficient of determination R^2^ = 0.50, i.e., ρ = 0.71), the required sample size was N = 16. This served as an approximate recruitment target rather than a formal power analysis for every reported endpoint. The 20 participants analyzed per protocol exceed that target.

Temporal correspondence between the two devices was assessed at the subject level using zero-lag Pearson correlation coefficients computed between the paired MCAv and Brain Flow index traces for each participant and protocol. The median coefficient was reported per protocol, and a one-sided Wilcoxon signed-rank test was used to assess whether the subject-level coefficients were greater than zero. The signals are dominated by the imposed block structure, so correlation magnitude is interpreted as temporal correspondence under the experimental perturbation rather than as an association between independent observations [43].

To account for possible temporal offsets between the two devices, we also computed lag-optimized correlations. For each participant and protocol, Pearson correlations were calculated across lags from -30 to +30 s in 1 s increments, and the lag yielding the maximum coefficient was retained; positive lags indicate that the Temple trace lagged behind the TCD trace, and negative lags that it led. This was done for MCAv and the Temple Brain Flow index. Median coefficients and their 95% confidence intervals were obtained by bootstrapping, and one-sided Wilcoxon signed-rank tests assessed whether the subject-level lag-optimized coefficients exceeded zero. Because the same traces are used both to select and to evaluate the best lag, the lag-optimized coefficients are reported alongside, and not in place of, the zero-lag result.

Cross-device heart-rate consistency was summarized as the subject-level mean absolute percentage error (MAPE) [44,45] between Temple-derived heart rate (Temple-HR) and TCD-derived heart rate (TCD-HR, the reference) across the full protocol:

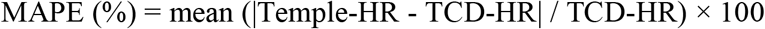

Transition responses were analyzed within each device by testing the null hypothesis that the median transition response was equal to zero (Wilcoxon signed-rank test); p-values were Bonferroni-corrected across all transition comparisons. These tests were applied to the distribution of individual subject transition responses. To visualize correspondence in relative transition magnitude, we also calculated paired differences after each full-session signal had been z-scored. The difference (MCAv minus Temple-BF) was plotted against the mean of the two standardized responses with mean ± 1.96 SD limits. The two modalities measure different physical quantities, and z-scoring imposes a common scale on them, so these plots are presented as standardized paired-difference displays and not as conventional evidence of interchangeable method agreement [46]. Within-session z-scoring rescales each trace to zero mean and unit standard deviation, so two signals sharing a response shape yield almost identical Δz regardless of their gain; this analysis is therefore sensitive to response shape and structurally insensitive to differences in response amplitude between the devices.

## 3. Results

Twenty participants contributed analyzable data to each protocol after protocol-specific quality exclusions. We first tested whether Temple-BF reproduced the expected direction and temporal structure of TCD-derived MCAv during exercise and postural change.

Figure 3 shows subject-averaged, z-scored traces of MCAv and the Brain Flow index. MCAv showed the expected temporal trends in both protocols: it increased during seated cycling and decreased during recovery (Figure 3a), and it was lower during standing and higher during supine epochs (Figure 3d). The Brain Flow index followed the same patterns in both protocols. Heart rate traces from the two systems were also closely aligned, but in the postural protocol heart rate decreased while MCAv and Temple-BF increased (Figure 3a and 3d, lower panels).

**Figure 3.**
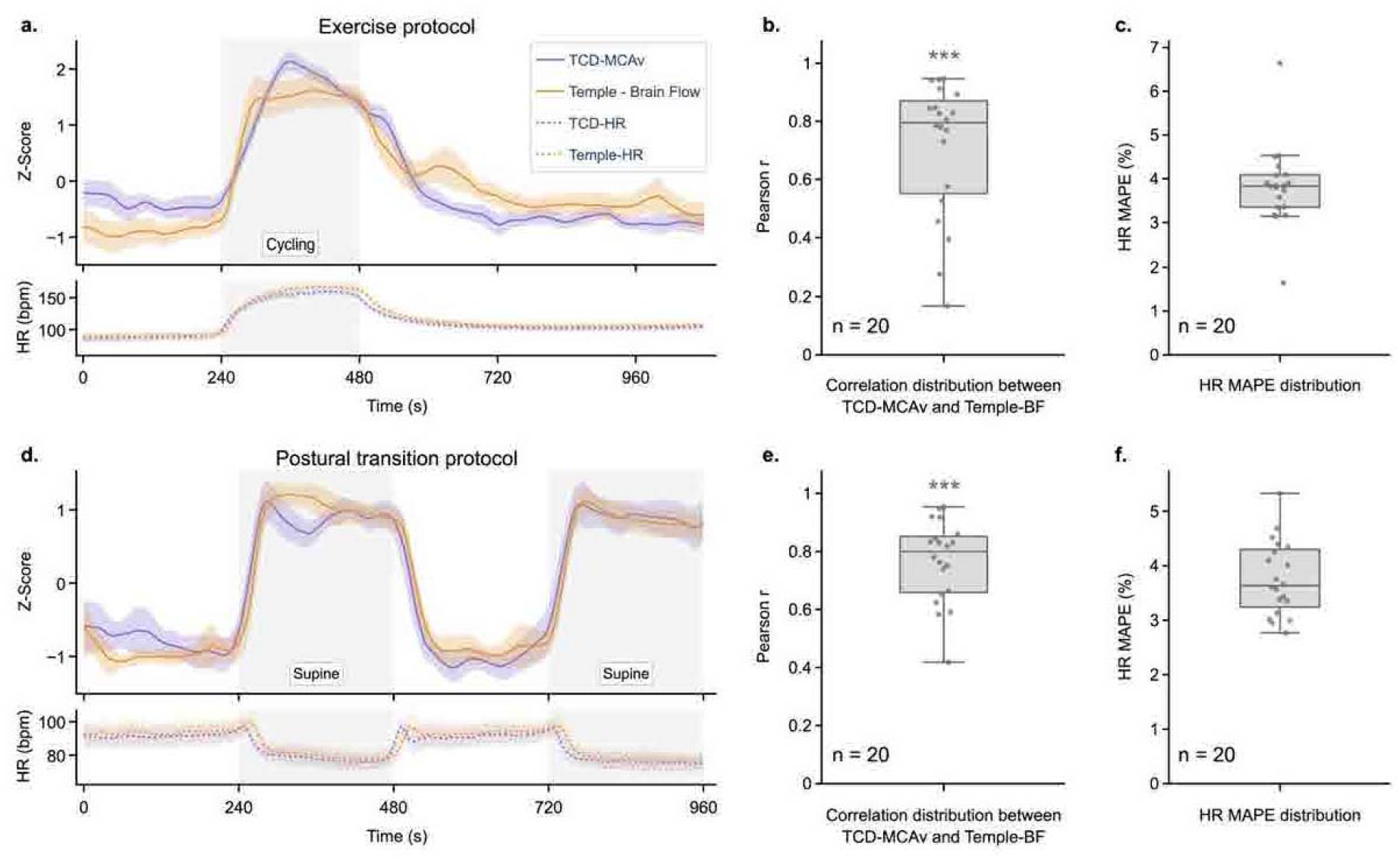
Subject-averaged traces and temporal correspondence. **(a, d)** Subject-averaged, z-scored time series (mean ± 95% CI) for TCD-derived MCAv (purple) and the Temple Brain Flow index (orange), with per-device heart rate below (dashed), for the **(a)** exercise and **(d)** postural transition protocols; shading marks the cycling and supine epochs. **(b, e)** Distributions of subject-level Pearson correlation coefficients between paired MCAv and Brain Flow index traces (median r = 0.795 exercise; r = 0.799 postural; one-sided Wilcoxon signed-rank test). **(c, f)** Distributions of subject-level heart rate mean absolute percentage error (HR MAPE). Asterisks (***) denote p < 0.001.

Subject-level zero-lag Pearson correlations between paired MCAv and Brain Flow index traces were positive (Figure 3b, e); the median coefficient was r = 0.795 in the exercise protocol and r = 0.799 in the postural transition protocol, and the distribution of subject-level coefficients was significantly greater than zero in both (one-sided Wilcoxon signed-rank test, both p < 0.001). These coefficients quantify the temporal correspondence between the two signals under the imposed protocol structure.

To establish whether accounting for temporal offset would improve the correlation, we calculated lagged correlations by shifting the two time series (see Methods). For the exercise protocol, the median lag-optimized correlation coefficient between MCAv and the Brain Flow index increased to 0.837 (95% CI, 0.718-0.884) at a median best lag of -19.0 s (95% CI, -26.5 to 5.0 s); for the postural transition protocol correlation coefficient was 0.817 (95% CI, 0.777-0.885) at a median lag of 9.0 s (95% CI, 0.0-16.0 s). This result indicated that the temporal ordering of the two traces depended on the protocol, with the Brain Flow index evolving faster than MCAv during exercise, whereas the opposite was true during the postural transition protocol. Both bootstrap intervals reach zero, however, and the two systems were aligned only by session timestamps (Section 2.2), so a protocol-dependent temporal ordering is suggested by these data but not established. Subject-level HR mean absolute percentage error (MAPE) remained low, with a median of 3.83% during the exercise protocol (interquartile range, IQR 3.35-4.08%, n = 20) and 3.63% during the postural transition protocol (IQR 3.30-4.28%, n = 20) (Figure 3c, f), indicating close agreement between Temple-HR and TCD-HR across participants and conditions.

Figure 4 shows transition-level responses. In the exercise protocol, MCAv increased from baseline to cycling and decreased from cycling to recovery, and the Brain Flow index followed the same direction (Figure 4a): the median baseline-to-cycling response was +2.12 Δz for MCAv and +2.46 Δz for the Brain Flow index, and the median cycling-to-recovery response was -2.26 Δz for MCAv and -1.84 Δz for the Brain Flow index. In the postural transition protocol, MCAv increased on standing-to-supine and decreased on supine-to-standing, and the Brain Flow index tracked the same pattern (Figure 4b): median responses were +1.85 Δz (MCAv) and +1.94 Δz (Brain Flow) for standing-to-supine, and -1.78 Δz (MCAv) and -2.03 Δz (Brain Flow) for supine-to-standing. All transition responses were significantly different from zero after Bonferroni correction (all adjusted p<0.001).

**Figure 4.**
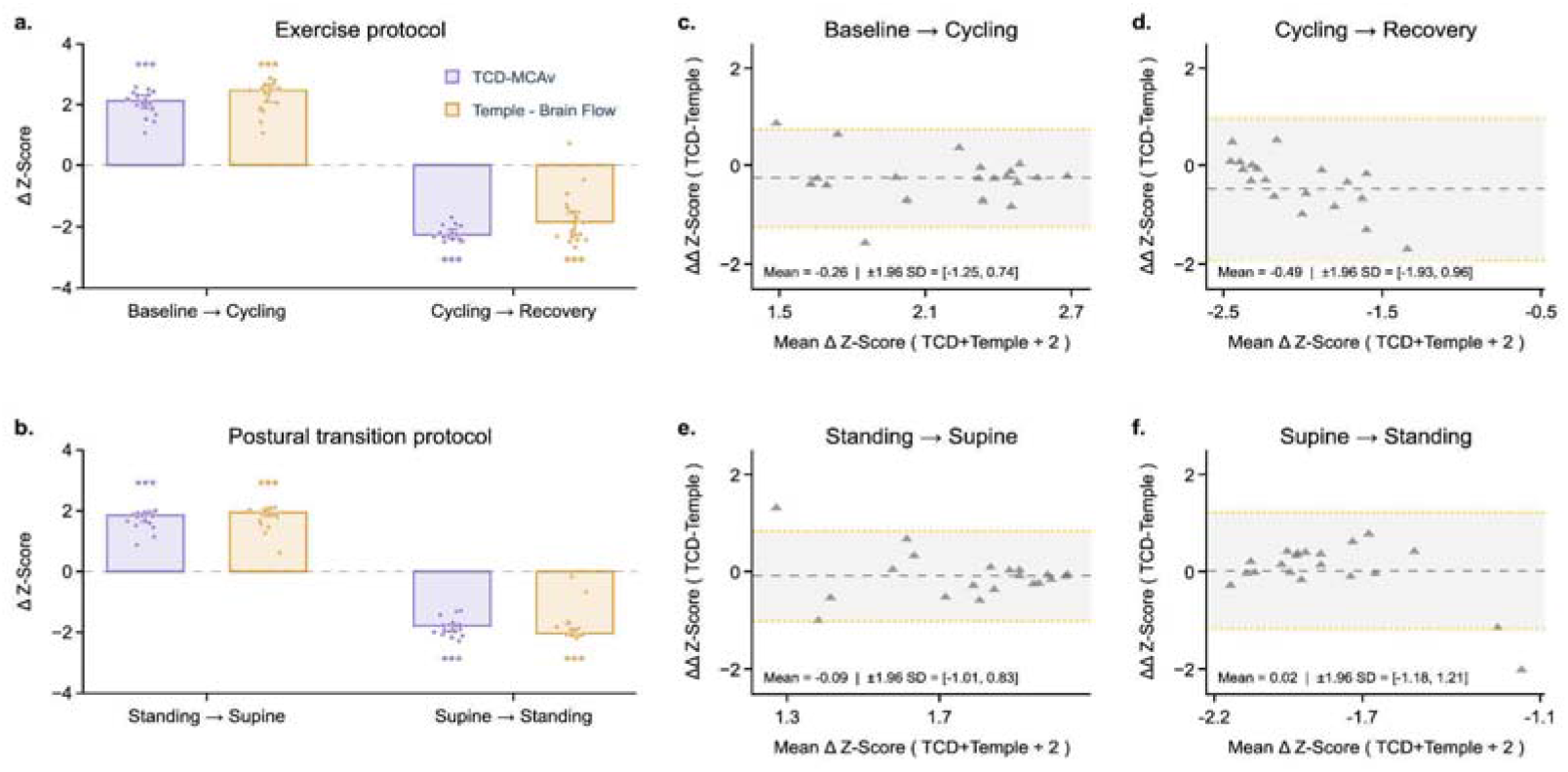
Transition-response magnitudes and Bland-Altman correspondence. **(a, b)** Group median transition magnitude (Δz-score) for TCD-derived MCAv (purple) and the Temple Brain Flow index (orange) in the **(a)** exercise and **(b)** postural transition protocol, with participant points overlaid; stars denote Wilcoxon signed-rank tests against zero with Bonferroni correction across eight comparisons (p < 0.001). **(c-f)** Bland-Altman plots for each transition: the inter-device difference (ΔΔz = MCAv - Brain Flow index) against the mean of the two device responses; each marker is one participant, the dashed line is the mean bias, and the shaded band is the 95% limits of agreement (mean ± 1.96 SD).

Next, we performed Bland-Altman analysis of the normalized transition responses (Figure 4c-f). The analysis was interpreted as an assessment of relative response agreement across transitions, rather than equivalence of the underlying device measurements. Because MCAv is measured in cm/s and the Brain Flow index is dimensionless, the limits of agreement are expressed in standardized units and have no direct interpretation in units of velocity. Mean biases were small: -0.26 Δz (baseline-to-cycling) and -0.49 Δz (cycling-to-recovery) in the exercise protocol, and - 0.09 Δz (standing-to-supine) and +0.02 Δz (supine-to-standing) in the postural transition protocol. These biases are small on the standardized scale, but that is largely a consequence of the normalization: z-scoring removes any difference in gain between the devices, so a near-zero bias indicates matched response shape rather than matched response amplitude. Inter-subject variability was nonetheless present, as expected given differences in measurement modality, anatomical sampling, and the physiological contributors to each signal. These analyses therefore describe how closely the two response profiles correspond, and should not be read as quantitative agreement in physical units or as evidence that Temple-BF and MCAv are interchangeable measurements.

## 4. Discussion

The principal finding is that Temple-BF covaried with TCD-derived MCAv across two physiologically distinct perturbations, with median zero-lag within-participant correlations close to 0.8 and concordant bidirectional transition responses. This result shows task-response concordance. The temporal correspondence indicates that the final wearable output contains dynamics that track MCAv under these conditions; additional studies are needed to indicate that the two signals share the same anatomical source or physical units.

The correspondence between the two signals is notable because they are obtained by different physical principles and at different depths. TCD measures blood-flow velocity within the middle cerebral artery, inside the cranial cavity, whereas the Temple device senses an optical signal at the surface of the anterior temple and therefore reflects extracranial as well as any deeper contributions. The most parsimonious interpretation is that the protocols used here, cycle-ergometer exercise and postural change, drive systemic, directionally shared changes in head blood flow, so a surface optical signal can track an intracranial velocity reference when both move together. Accordingly, the present findings support the use of the Brain Flow index as a relative marker of hemodynamic change during whole-head physiological perturbations.

The MCAv responses observed here were consistent with previously reported cerebrovascular responses to cycle-ergometer exercise [28,30] and to postural change [6,7,31]. These expected directional changes support the physiological validity of the protocols and provide a reference pattern against which the device output could be evaluated. The clear and reproducible difference in MCAv between standing and supine is itself consistent with the contemporary understanding that the cerebral autoregulatory range is narrower than the classical model implied, such that ordinary postural shifts in perfusion pressure produce measurable changes in cerebral blood flow [33,34].

The device also tracked heart rate well, with a mean absolute percentage error (MAPE) of approximately 4% during cycle-ergometer exercise. This provides an independent check that the sensor and its pulse-detection pipeline behaved as expected during movement and indicates that the temporal offsets between the two cerebrovascular signals reflect genuine physiological and measurement differences rather than sensor failure. The two instruments showed small temporal offsets whose direction differed between protocols, so a fixed delay in either signal does not account for them. Differences in response kinetics between an arterial velocity measurement and a tissue-level optical composite, smoothing within the Brain Flow algorithm, alignment of two independently clocked systems, and the different pressures exerted on the head and the device during each protocol could each contribute. These contributions can be studied in future work.

A systems view helps reconcile the strong temporal associations between MCAv and Temple-BF. Dynamic cerebral autoregulation is conventionally treated as an input-output relation in which fluctuations in MAP are transformed into changes in cerebral flow or velocity, with gain, phase, and coherence describing the frequency-dependent response [24,25]. Cutaneous vascular beds have their own pressure-flow dynamics, which differ between skin types; this has been characterised for glabrous and non-glabrous skin of the palm and forearm rather than for facial skin [47]. Temple-BF may thus represent an empirical transformation of local PPG-derived information that is sensitive to the same systemic forcing that reaches the cerebral circulation. The present data cannot determine whether the algorithm preferentially retains cerebral-like pressure-response dynamics, because beat-to-beat pressure and local temple perfusion were not measured.

### 4.1. Interpretation in the context of head-worn optical wearables

Temple-BF belongs to a family of head-surface optical measurements that includes photoplethysmography, near-infrared spectroscopy (NIRS), and diffuse correlation methods. The forehead and temple are practical sites for wearable optical sensing because of their thin skin, dense superficial vasculature, and relative resistance to motion artifact and peripheral vasoconstriction compared with the fingertip [48,49]. Forehead photoplethysmography has shown systematic perfusion changes during head-down tilt [50]. More broadly, wearables that interrogate external carotid territories have been proposed as indirect indices of carotid or cerebral blood flow change; for example, an in-ear sensor over the posterior auricular artery tracked carotid blood flow changes and preceded syncope during tilt-table testing [51]. Together with the present findings, these studies support the feasibility of head-surface optical sensing for tracking physiologically meaningful perfusion changes.

However, these modalities do not measure the same physical quantity as TCD. MCAv reflects blood flow velocity in a large intracranial artery, whereas optical signals reflect a composite of superficial and deep tissue hemodynamics, including changes in hemoglobin concentration and oxygenation [20]. Temple-BF was therefore evaluated against MCAv as a relative signal rather than as a direct measure of cerebral blood flow.

The present findings support this response-level interpretation. During exercise, heart rate and MCAv increased together. In contrast, during postural transitions, MCAv increased on returning to supine despite a lower heart rate. This provides a useful discriminating test: a signal driven solely by heart rate would be expected to change in the opposite direction. Instead, the Brain Flow index tracked MCAv across both protocols, indicating that it is not simply a surrogate for heart rate.

The observed correspondence should be interpreted as complementary to, rather than interchangeable with, TCD. The postural transitions used here are likely to influence intracranial and extracranial perfusion in the same direction; thus, correspondence with MCAv is expected under these conditions and does not by itself establish a specifically cerebral contribution. Establishing the cerebral specificity and clinical utility of the Temple-BF signal will require studies in broader populations, with complementary depth-resolved optical measurements and protocols specific to cerebral hemodynamic changes, such as hypercapnia [52] or lower-body negative pressure [53].

### 4.2. Limitations and Future Directions

Several considerations bound the interpretation of these findings. TCD-derived MCAv reflects blood-flow velocity in a single, unilateral artery and indexes cerebral blood flow under the assumption of an approximately constant insonated diameter [54,55], so it is best regarded as a well-validated surrogate for, rather than a direct measure of, volumetric cerebral blood flow. The Brain Flow index, for its part, is an extracranial optical signal whose cerebral component, as set out above, the present protocols cannot isolate. Agreement was quantified as directional concordance and relative temporal correspondence, an approach well suited to demonstrating that the two signals covary, though not to certifying quantitative accuracy against absolute cerebral blood flow. The Bland-Altman analysis is subject to the same constraint. It was performed on within-session standardized responses rather than on values in shared physical units, so it tests whether the two signals report the same relative change, and its limits of agreement carry no interpretation in cm/s. The standing-to-supine response also averaged two transitions whereas the supine-to-standing response was derived from a single transition, so the two posture panels are not estimated with equal precision. Finally, the index is generated by a proprietary algorithm that cannot be inspected, the source-detector separation and detector geometry were not disclosed, and the cohort comprises healthy young adults studied under controlled conditions in modest numbers. Temple and TCD were recorded over contralateral temples, so the two devices did not sample the same hemisphere.

One element of consistency is already available within the present design. The postural protocol repeated the standing and supine blocks, so each participant contributed two standing-to-supine transitions; the reported response averages the two. Comparing the cycles separately would test within-session consistency directly. Reliability across separate visits was not assessed here. These considerations do not diminish the present correspondence, but they outline a clear path for building on it. Larger and more diverse cohorts, assessment of long-term wear and day-to-day repeatability, and additional targeted challenges such as paced breathing, alongside the decoupling protocols described above, would each sharpen the interpretation of Temple-BF. Simultaneous depth-resolved functional near-infrared spectroscopy (fNIRS) measurements could further help establish the metric’s cerebral specificity.

## 5. Conclusions

Across a cycle-ergometer exercise protocol and a stand-to-supine postural transition protocol in healthy adults, a temple-worn optical wearable’s Brain Flow index tracked TCD-derived MCAv, with significant subject-level temporal correlations and directionally concordant transition responses. Within the postural protocol, MCAv and Temple-BF both increased in the supine position while heart rate decreased, which is evidence against a simple heart-rate artifact, although pressure, CO2, hydrostatic, autonomic, and extracranial explanations all remain open. These findings show that the Brain Flow index tracks a cerebrovascular reference under protocols of this kind, on a relative rather than an absolute scale, and that it does not substitute for TCD-measured cerebral blood-flow velocity. Further studies in broader populations, using complementary modalities and targeted cerebrovascular reactivity protocols, are needed to establish the cerebral specificity of the Temple-BF signal and define when and for whom it is informative.

## Author Contributions

Conceptualization, S.P. and N.J.G. and L.V.R.; methodology, and relevant permission, N.J.G.; human resources and clinical supervision, R.C.G.; data acquisition,A.K. and S.K.S.; curation of device-derived data and formal analysis, A.G. and N.J.G.; supervision, visualization, and original draft preparation, A.K., L.V.R. and N.J.G.; review and editing, A.G., R.C.G and S.P. All authors have read and agreed to the published version of the manuscript.

## Funding

Financial assistance was granted to N.J.G. at Chaudhary Charan Singh University by Temple Pvt. Ltd. (Gurugram, India).

## Conflicts of Interest

N.J.G. received research funding from Temple Pvt. Ltd. for this work (see Funding). S.P. is a paid scientific consultant to Temple Pvt. Ltd. The other listed authors declare no personal conflict of interest. Temple Pvt. Ltd. provided the wearable devices and returned de-identified device-derived outputs for the requested session timestamps. The academic investigators retained responsibility for the TCD measurements and study analyses reported here.

## Institutional Review Board Statement

The study was conducted in accordance with the Declaration of Helsinki and approved by the Institutional Human Ethics Committee of Lala Lajpat Rai Memorial Medical College, Chaudhary Charan Singh University, Meerut, India (approval number SC-1/2025/9370; date of approval 16 December 2025). All procedures followed the ethical guidelines of the Department of Health Research (DHR), Indian Council of Medical Research (ICMR), India.

## Informed Consent Statement

Informed consent was obtained from all subjects involved in the study.

## Data Availability Statement

The de-identified summary data and analysis code that support the findings of this study are available from the corresponding author upon reasonable request.

## Acknowledgments

The authors thank Temple Pvt. Ltd. for providing three Temple devices, usage guidance, backend app data with timestamps, and insights into the device-derived data. We thank anonymous participants, who volunteered for the study.

## Notes

### Author Declarations

Institutional Human Ethics Committee of Lala Lajpat Rai Memorial Medical College, Chaudhary Charan Singh University, Meerut, India gave approval for this work vide approval number SC-1/2025/9370 dated 16 December 2025 as per ethical guidelines of the Department of Health Research (DHR), Indian Council of Medical Research (ICMR), India.

### Summary of Updates

clarified competing interest statement to match the same in the manuscript

